# Associations Between Stroke Type, Ischemic Stroke Subtypes, and Post-Stroke Cognitive Trajectories

**DOI:** 10.1101/2024.04.29.24306600

**Authors:** Deborah A. Levine, Rachael T. Whitney, Wen Ye, Emily M. Briceño, Alden L. Gross, Bruno J. Giordani, Jeremy B. Sussman, Ronald M. Lazar, Virginia J. Howard, Hugo J. Aparicio, Alexa S. Beiser, Mitchell S. V. Elkind, Rebecca F. Gottesman, Silvia Koton, Sarah T. Pendlebury, Adam S. Kollipara, Mellanie V. Springer, Sudha Seshadri, Jose R. Romero, Annette L. Fitzpatrick, William T. Longstreth, Rodney A. Hayward

**Affiliations:** Department of Internal Medicine, University of Michigan (U-M), Ann Arbor, MI; Department of Neurology, U-M, Ann Arbor, MI; Department of Biostatistics, U-M, Ann Arbor, MI; Department of Physical Medicine and Rehabilitation, U-M, Ann Arbor, MI; Department of Epidemiology, Johns Hopkins University, Baltimore, MD; Department of Psychiatry, U-M, Ann Arbor, MI; VA Ann Arbor Healthcare System, Ann Arbor, MI; Department of Neurology University of Alabama at Birmingham, AL; Department of Epidemiology, University of Alabama at Birmingham, AL; Department of Neurology, Boston University, MA; Department of Biostatistics, Boston University, MA; Department of Neurology, Columbia University, New York, NY; Stroke Branch, National Institute of Neurological Disorders and Stroke, Bethesda, MD; Department of Nursing, Tel Aviv University, Israel; Nuffield Department of Clinical Neurosciences, University of Oxford, UK; NIHR Biomedical Research Centre, Departments of Medicine and Geratology, Oxford University Hospitals NHS Foundation Trust, UK; Department of Neurology, University of Texas San Antonio, TX; Department of Epidemiology, University of Washington, Seattle, WA

**Keywords:** Cognition, Stroke, Blood Pressure, Glucose, Cholesterol

## Abstract

**Background:** It is unclear how post-stroke cognitive trajectories differ by stroke type and ischemic stroke subtype. We studied associations between stroke types (ischemic, hemorrhagic), ischemic stroke subtypes (cardioembolic, large artery atherosclerotic, lacunar/small vessel, cryptogenic/other determined etiology), and post-stroke cognitive decline.

**Methods:** This pooled cohort analysis from four US cohort studies (1971-2019) identified 1,143 dementia-free individuals with acute stroke during follow-up: 1,061 (92.8%) ischemic, 82 (7.2%) hemorrhagic, 49.9% female, 30.8% Black. Median age at stroke was 74.1 (IQR, 68.6, 79.3) years. Outcomes were change in global cognition (primary) and changes in executive function and memory (secondary). Outcomes were standardized as T-scores (mean [SD], 50 [10]); a 1-point difference represents a 0.1-SD difference in cognition. Median follow-up for the primary outcome was 6.0 (IQR, 3.2, 9.2) years. Linear mixed-effects models estimated changes in cognition after stroke.

**Results:** On average, the initial post-stroke global cognition score was 50.78 points (95% CI, 49.52, 52.03) in ischemic stroke survivors and did not differ in hemorrhagic stroke survivors (difference, -0.17 points [95% CI, -1.64, 1.30]; *P*=0.82) after adjusting for demographics and pre-stroke cognition. On average, ischemic stroke survivors showed declines in global cognition, executive function, and memory. Post-stroke declines in global cognition, executive function, and memory did not differ between hemorrhagic and ischemic stroke survivors. 955 ischemic strokes had subtypes: 200 (20.9%) cardioembolic, 77 (8.1%) large artery atherosclerotic, 207 (21.7%) lacunar/small vessel, 471 (49.3%) cryptogenic/other determined etiology. On average, small vessel stroke survivors showed declines in global cognition and memory, but not executive function. Initial post-stroke cognitive scores and cognitive declines did not differ between small vessel survivors and survivors of other ischemic stroke subtypes. Post-stroke vascular risk factor levels did not attenuate associations.

**Conclusion:** Stroke survivors had cognitive decline in multiple domains. Declines did not differ by stroke type or ischemic stroke subtype.

## INTRODUCTION

Incident stroke is associated with accelerated, yearslong cognitive decline^1^ and dementia.^2^ Most strokes are ischemic (85-90%); fewer are intracerebral hemorrhages (10%) and subarachnoid hemorrhages (3%). Ischemic strokes can be classified as cardioembolic (∼20%), large-artery atherosclerotic (∼15%), lacunar/small vessel (∼20-25%), cryptogenic (∼30-40%), and other determined etiology (<5%).^3^ Relationships between stroke type, ischemic stroke subtype, and post-stroke cognitive decline and dementia are unclear; some studies found greater risk for hemorrhagic and cardioembolic strokes.^4,5^ How vascular risk factor (VRF) levels affect these relationships is also unclear. One study suggested that VRF lowering might reduce post-stroke dementia risk in atherosclerotic ischemic strokes, but not hemorrhagic or cardioembolic ischemic strokes.^6^

Understanding relationships between stroke type, ischemic stroke subtype, and VRF levels with cognitive decline will suggest potential interventions and high-risk groups to target. Sufficient sample size and pre-stroke cognition are required to assess these relationships meaningfully. We analyzed pooled individual participant data from four cohort studies—with expert-adjudicated incident strokes, stroke subtyping, and repeated objective measures of cognition pre- and post-stroke—to clarify relationships between stroke type, ischemic stroke subtype, and post-stroke cognitive trajectories, and to explore how VRFs affect these relationships. We hypothesized that all stroke types and ischemic stroke subtypes are associated with post-stroke cognitive decline, and that post-stroke systolic blood pressure (SBP), glucose, and lipid levels partially explain these associations.

## METHODS

Data are available from the corresponding author upon cohort and research team approval, and data use agreements with cohorts and the corresponding author’s institution.

### Study Design, Participants, and Measurements

STROKE COG pooled participant data from cohort enrollment through December 31, 2019 from four US cohort studies: Atherosclerosis Risk in Communities (ARIC) Study,^7^ Cardiovascular Health Study (CHS),^8^ Framingham Offspring Study (FOS),^9^ and Reasons for Geographic and Racial Differences in Stroke (REGARDS) Study.^10^ These cohorts had ≥50 participants with physician-adjudicated acute stroke during follow-up and SBP and cognition measures pre- and post-stroke.^11^

We included participants ≥18 years with expert-adjudicated acute ischemic or hemorrhagic stroke during follow-up, ≥1 pre-stroke and ≥1 post-stroke cognitive assessments, and ≥1 post-stroke SBP measurement at or before the last post-stroke cognitive assessment. We excluded participants with cohort-defined dementia at or before stroke and those reporting Hispanic ethnicity or race other than Black or White due to cohort design differences. We included participants with self-reported history of stroke at cohort baseline in the primary analysis as their inclusion did not change estimates of an acute stroke’s effect on cognitive trajectories controlling for pre-stroke cognition.^1,11^ We performed a sensitivity analysis excluding participants with a self-reported history of stroke at cohort baseline.

The University of Michigan Institutional Review Board approved this study. Participating institutions’ review boards approved the cohort studies. All participants provided written informed consent.

### Stroke Measurement

Expert physicians adjudicated stroke cases using medical records, proxy interviews, and/or death certificates and comparable clinical criteria.^12-15^ Expert physicians classified strokes as hemorrhagic (intracerebral/subarachnoid) or ischemic with ischemic subtype.^16^ We restricted hemorrhagic strokes to intracerebral hemorrhage except for CHS, which combined subarachnoid and intracerebral hemorrhage. Ischemic strokes were classified as cardioembolic, large artery/carotid atherosclerotic, lacunar/small vessel, and cryptogenic/other determined etiology.^16^ We combined cryptogenic/other determined etiology because ARIC did not distinguish between them.

### Cognitive Function

Cognitive tests consistent with the Vascular Cognitive Impairment Harmonization Standards^17^ were administered to participants in-person (ARIC, CHS, FOS) or by telephone (REGARDS) using standardized protocols. Global cognition, executive function, and memory can be measured reliably and validly over the telephone.^18^ We co-calibrated available cognitive test items across cohorts into three domains representing global cognition, memory, and executive function using confirmatory factor analysis and item response theory (**eMethods**).^19-21^ Factor scores were estimated in Mplus^22^ and set to a T-score metric (mean 50, standard deviation [SD] 10) at each participant’s first post-stroke cognitive assessment. A 1-point difference in factor score represents a 0.1 SD difference in the distribution of cognitive function across cohorts. Higher scores indicated better performance. Outcomes were change in global cognition (primary) and changes in executive function and memory domains (secondary).

### Post-Stroke Vascular Risk Factors

Cohorts measured SBP, glucose, and low-density lipoprotein (LDL) cholesterol at each in-person exam using standard protocols and equipment. We summarized VRFs as time-dependent cumulative means of all post-stroke measurements at or before each post-stroke cognitive assessment.^23^

### Covariates

Covariate selection was based on a conceptual model^5^, cohort availability, and prior research^11^ (**eMethods**). Covariates were age, self-identified sex, self-identified race, education, cohort, number of apolipoprotein E (ApoE) ε4 alleles, and depressive symptoms measured using the Centers for Epidemiologic Studies Depression (CES-D) Scale. Race is a social—not biological—construct used to gauge our sample’s diversity. Pre-stroke global cognition, executive function, and memory scores were summarized as the time-invariant mean of all pre-stroke measurements.

### Statistical Analysis

We used linear mixed-effects models to estimate longitudinal change in each continuous cognitive outcome by stroke type and ischemic stroke subtype. Ischemic stroke was the reference group. Analyses of ischemic stroke subtype were restricted to ischemic stroke survivors from ARIC, CHS, and REGARDS because FOS only measured small vessel and cardioembolic subtype. Small vessel was the reference group. We imputed education for nine participants using multiple imputation.

**Model 1** included follow-up time, stroke subtype × follow-up time, age × follow-up time, sex, race, education, cohort, mean pre-stroke cognition, and subject-specific random effects for intercept and slope. Follow-up time was defined as years since stroke and treated as a continuous variable. **Model 2** added time-varying cumulative mean post-stroke SBP, glucose, and LDL cholesterol, and their respective interactions with follow-up time to **Model 1**. Statistical significance for all analyses was set as *P*<0.05 (2-sided). There was no evidence of race × follow-up time (*P*=0.32) or sex × follow-up time (*P*=0.10) interactions in analyses of stroke type and the primary outcome.

Analyses were performed using Stata version 18.0 (StataCorp).

### Sensitivity Analyses

To evaluate the robustness of our findings, we repeated analyses: 1) including CES-D score and CES-D score × time in the subset with CES-D data; 2) including ApoE genotype and ApoE genotype × time in the subset with genetic data; 3) excluding participants with self-reported history of stroke at cohort baseline; and 4) censoring observations at time of second incident stroke.

### Access to Data

Drs. Levine and Whitney had full access to all study data and take responsibility for the integrity of the data and analysis.

## RESULTS

Figure 1 presents a STROBE diagram. The final sample included 1,143 dementia-free participants with acute stroke during follow-up: 1,061 (92.8%) ischemic, 82 (7.2%) hemorrhagic, 570 (49.9%) female, 352 (30.8%) Black. Median age at incident stroke was 74.1 (IQR, 68.6, 79.3) years and the median follow-up time after stroke for the primary outcome was 6.0 (IQR, 3.2, 9.2) years. **Table 1** presents participant characteristics by stroke type. Median follow-up time after stroke for global cognition was 5.8 (IQR, 3.1, 9.2) for ischemic stroke survivors and 6.9 (IQR, 3.9, 9.5) for hemorrhagic stroke survivors (*P*=0.30).

**Figure 1.**
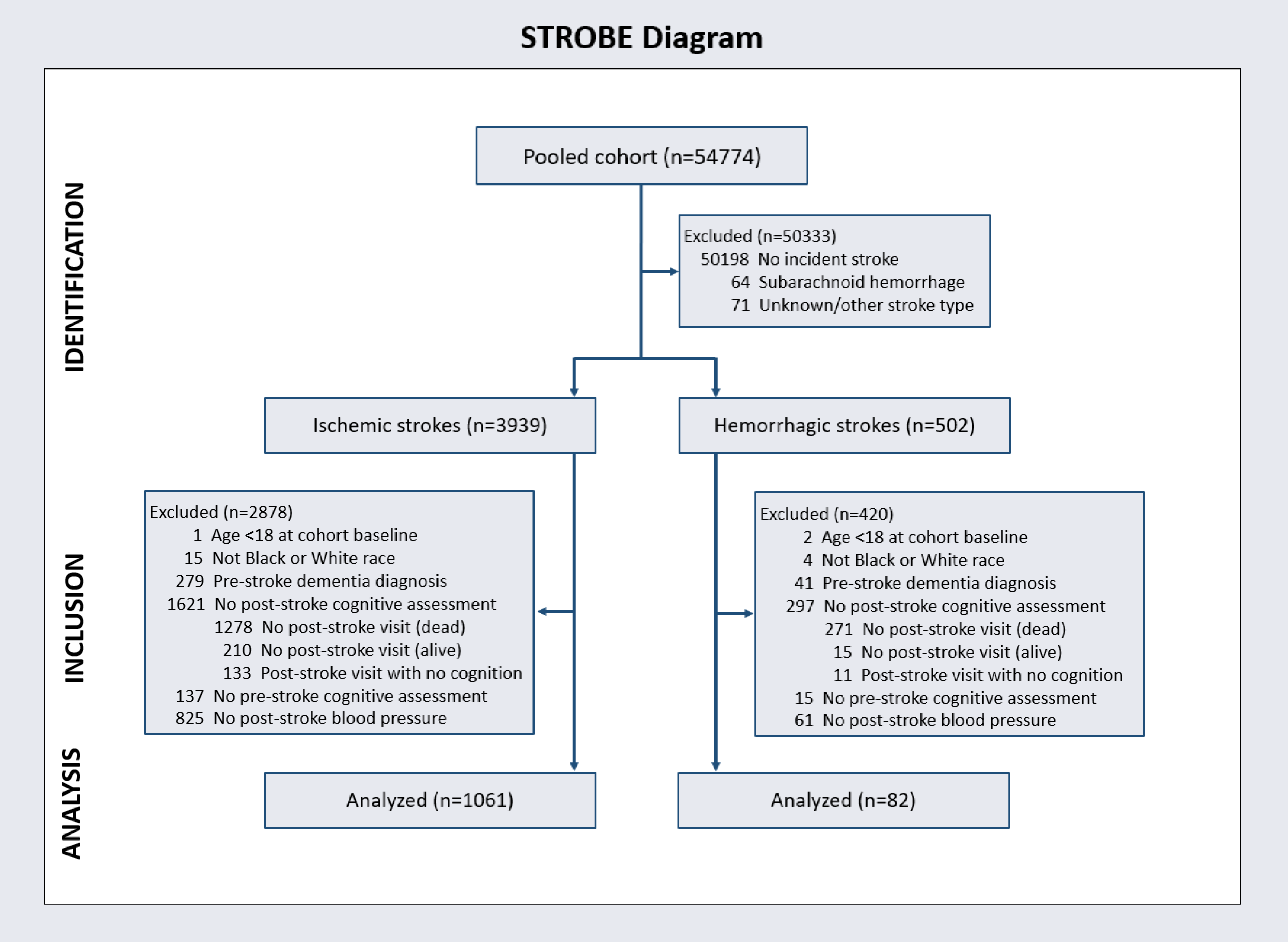
Derivation of the participant cohort.

**Table 1.** Characteristics of participants at first post-stroke cognitive assessment, by stroke type.

| Characteristic | Stroke Type (n=1,143) |  | P |
| --- | --- | --- | --- |
|  | Ischemic<br>(n=1,061) | Hemorrhagic<br>(n=82) |  |
| Age at stroke, y, range | 45.5–96.4 | 53.0–89.6 |  |
| Age at stroke, y, median (IQR) | 74.1 (68.8, 79.2) | 73.3 (67.0, 79.8) | 0.54 |
| Women, No. (%) | 529 (49.9) | 41 (50.0) | 0.98 |
| Race, No. (%) |  |  | 0.05 |
| White | 742 (69.9) | 49 (59.8) |  |
| Black | 319 (30.1) | 33 (40.2) |  |
| Cohort, No. (%) |  |  | 0.03 |
| ARIC | 229 (21.6) | 17 (20.7) |  |
| CHS | 324 (30.5) | 16 (19.5) |  |
| FOS | 105 (9.9) | 5 (6.1) |  |
| REGARDS | 403 (38.0) | 44 (53.7) |  |
| Education, No. (%) |  |  | 0.10 |
| Less than high school | 196 (18.6) | 8 (9.9) |  |
| Completed high school | 300 (28.5) | 27 (33.3) |  |
| Some college, no degree | 256 (24.3) | 16 (19.8) |  |
| College graduate or more | 301 (28.6) | 30 (37.0) |  |
| Ischemic stroke subtype, No. (%) |  |  |  |
| Cardioembolic | 200 (18.8) |  |  |
| Large artery atherosclerotic | 77 (7.3) |  |  |
| Lacunar/small vessel | 207 (19.5) |  |  |
| Cryptogenic/other determined etiology | 471 (44.4) |  |  |
| Missing | 106 (10.0) |  |  |
| Follow-up time after stroke, global cognition, y, median (IQR) | 5.8 (3.1, 9.2) | 6.9 (4.2, 9.5) | 0.30 |
| Follow-up time after stroke, executive function, y, median (IQR) | 5.4 (2.8, 8.6) | 6.2 (3.2, 8.9) | 0.46 |
| Follow-up time after stroke, memory, y, median (IQR) | 5.4 (2.6, 8.4) | 6.9 (3.9, 9.6) | 0.045 |
| SBP at first post-stroke cognitive assessment, mmHg, mean (SD) | 138.9 (23.8) | 138.2 (20.4) | 0.85 |
| No. SBP measurements after stroke, median (IQR) | 1.0 (1.0, 2.0) | 1.0 (1.0, 2.0) | 0.70 |
| Time from stroke to first post-stroke SBP, y, median (IQR) | 2.0 (0.7, 4.4) | 2.3 (0.7, 4.5) | 0.76 |
| Glucose at first post-stroke cognition, mg/dL, mean (SD) | 111.7 (37.4) | 105.8 (12.8) | 0.47 |
| No. glucose measurements after stroke, median (IQR) | 1.0 (1.0, 1.0) | 1.0 (1.0, 1.0) | 0.25 |
| Time from stroke to first post-stroke glucose, y, median (IQR) | 2.5 (1.1, 4.7) | 2.8 (1.0, 4.8) | 0.78 |
| LDL cholesterol at first post-stroke cognition, mg/dL, mean (SD) | 99.5 (36.6) | 109.9 (33.3) | 0.15 |
| No. LDL cholesterol measurements after stroke, median (IQR) | 1.0 (1.0, 1.0) | 1.0 (1.0, 1.0) | 0.09 |
| Time from stroke to first post-stroke LDL cholesterol, y, median (IQR) | 2.8 (1.3, 5.4) | 3.0 (1.0, 4.8) | 0.41 |
| **Pre-stroke cumulative mean cognitive scores** |  |  |  |
| Global cognition, median (IQR) | 52.6 (47.9, 56.5) | 53.5 (49.6, 57.1) | 0.14 |
| Executive function, median (IQR) | 49.9 (43.5, 54.8) | 51.6 (44.8, 57.2) | 0.16 |
| Memory, median (IQR) | 53.8 (50.0, 57.4) | 55.3 (52.1, 57.4) | 0.01 |
| **No. pre-stroke cognitive assessments** |  |  |  |
| Global cognition, median (IQR) | 4.0 (2.0, 7.0) | 4.0 (2.0, 6.0) | 0.85 |
| Executive function, median (IQR) | 2.0 (1.0, 4.0) | 2.0 (1.0, 3.0) | 0.01 |
| Memory, median (IQR) | 3.0 (2.0, 6.0) | 3.0 (2.0, 5.0) | 0.75 |
| **Cognitive scores at first post-stroke assessment** |  |  |  |
| Global cognition, median (IQR) | 49.8 (40.4, 57.8) | 50.2 (42.6, 58.3) | 0.46 |
| Executive function, median (IQR) | 44.9 (37.9, 52.4) | 47.2 (40.8, 52.8) | 0.17 |
| Memory, median (IQR) | 49.3 (43.5, 57.4) | 49.2 (49.1, 57.4) | 0.59 |
| **No. post-stroke cognitive assessments** |  |  |  |
| Global cognition, median (IQR) | 4.0 (2.0, 9.0) | 7.0 (2.0, 12.0) | 0.01 |
| Executive function, median (IQR) | 3.0 (2.0, 4.0) | 3.0 (2.0, 4.0) | 0.24 |
| Memory, median (IQR) | 3.0 (1.0, 6.0) | 5.0 (2.0, 8.0) | 0.006 |

SBP, systolic blood pressure; LDL, low-density lipoprotein.

Ischemic stroke survivors had fewer post-stroke global cognition (median [IQR], 4 [2, 9] vs. 7 [2, 12]; *P*=0.01) and memory (median [IQR], 3 [1, 6] vs. 5 [2, 8]; *P*=0.006) assessments than hemorrhagic stroke survivors. Of 1,061 ischemic strokes, 200 (18.8%) were cardioembolic, 77 (7.3%) large artery atherosclerotic, 207 (19.5%) lacunar/small vessel, 471 (44.4%) cryptogenic/other determined etiology, and 106 (10%) were missing classification. After excluding the 106 with missing classification, the ischemic stroke subtype analysis had 955 participants.

### Change in Global Cognition

On average, initial post-stroke global cognition score was 50.78 points (95% CI, 49.52, 52.03) in ischemic stroke survivors and did not differ in hemorrhagic stroke survivors (difference, -0.17 [95% CI, -1.64, 1.30] points; *P*=0.82) after adjusting for demographics and mean pre-stroke cognition (**Table 2**, **Model 1**). On average, ischemic stroke survivors experienced global cognition decline over time (-0.35 [95% CI, -0.43, -0.27] points/year; *P*<0.001) (**Table 2**, **Model 1**). Post-stroke global cognitive decline did not differ between hemorrhagic and ischemic stroke survivors. No consistent evidence showed that cumulative mean post-stroke SBP, glucose, or LDL cholesterol attenuated the association between stroke type and post-stroke global cognition (**Table 2**, **Model 2**). Older age was associated with faster post-stroke cognitive decline.

**Table 2.**
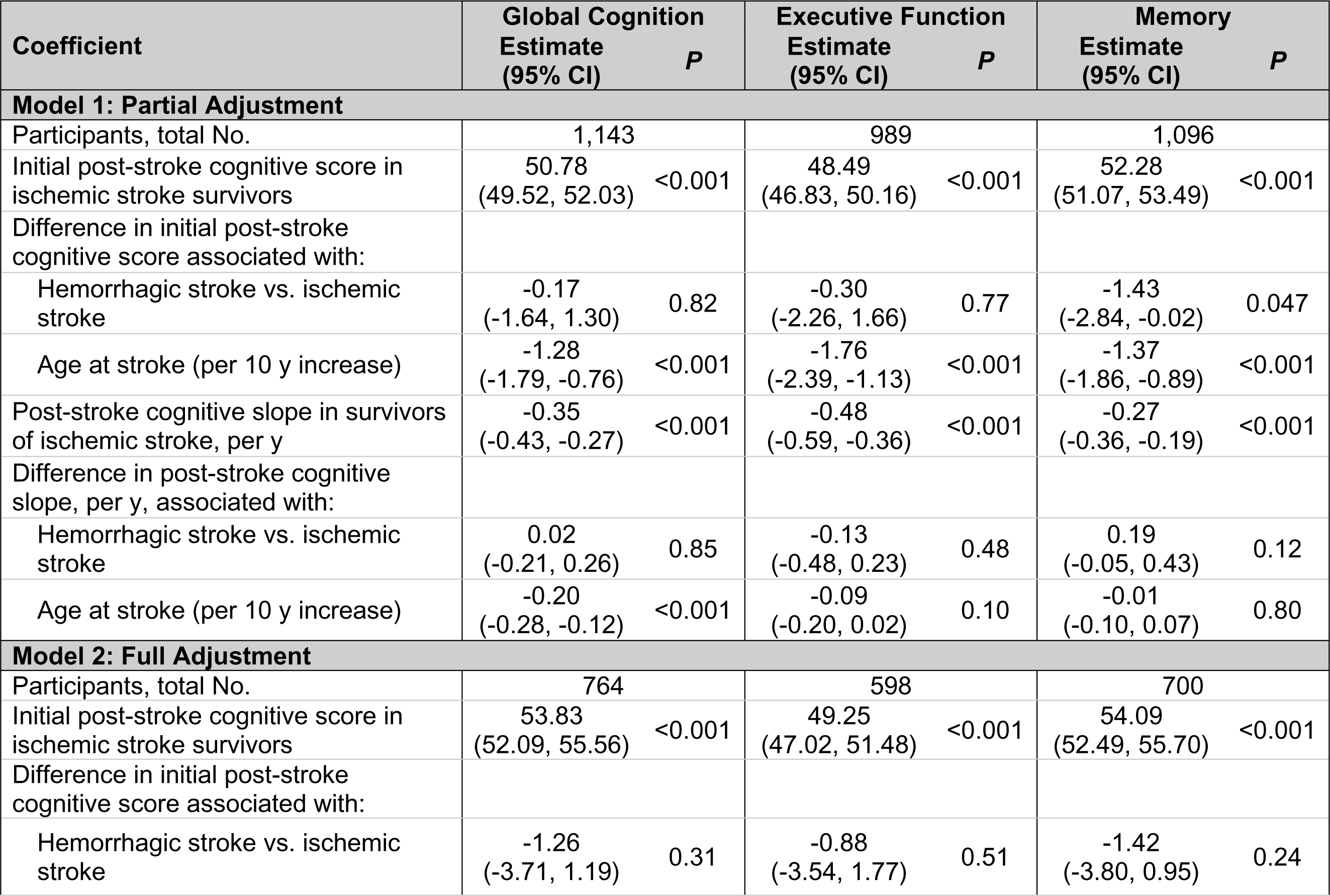
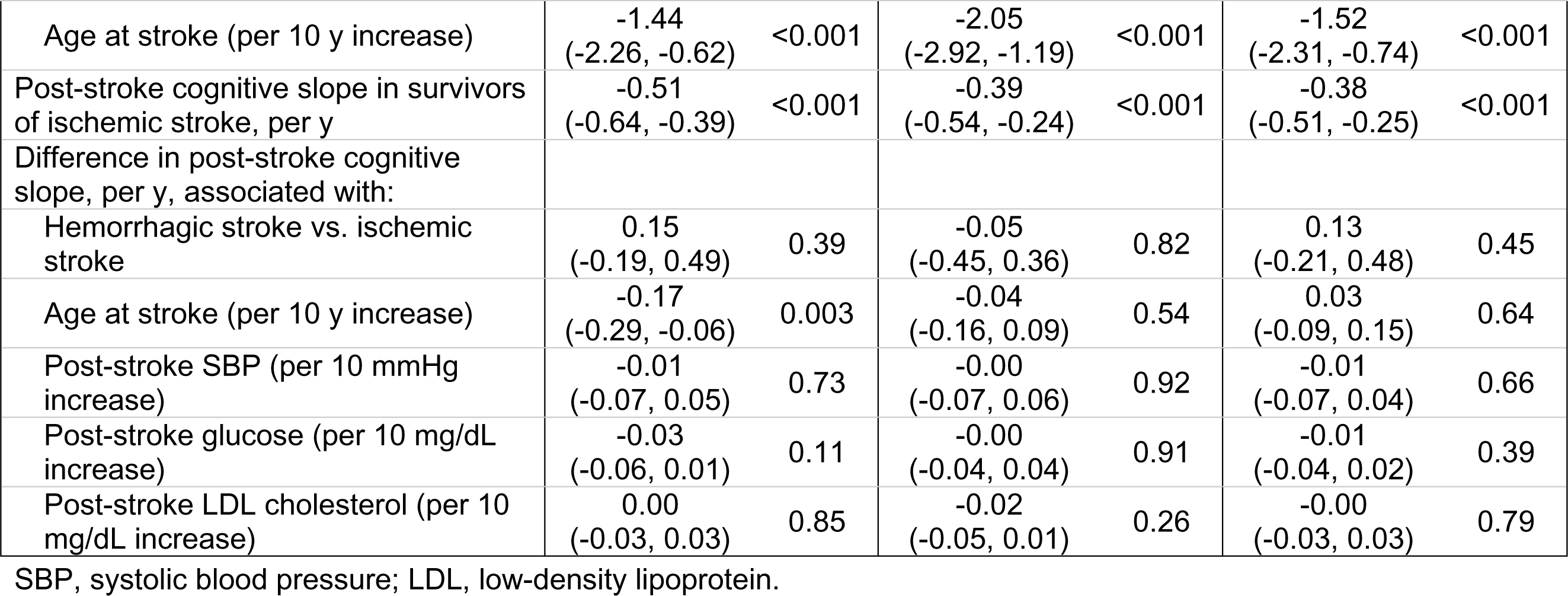
Association of stroke type and post-stroke cognitive decline.

In the subset with non-missing ischemic stroke subtype, on average, initial post-stroke global cognition score was 50.82 (95% CI, 49.28, 52.36) in small vessel survivors and did not differ in survivors of cardioembolic stroke, large artery atherosclerotic stroke, or cryptogenic/other determined etiology stroke after adjusting for age, sex, education, cohort, and mean pre-stroke cognition (**Table 3**, **Model 1**). On average, small vessel survivors experienced global cognition decline over time (-0.33 [95% CI, -0.49, -0.16] points/year; *P*<0.001) (**Table 3**, **Model 1**). Global cognitive decline after stroke did not differ between small vessel survivors and survivors of cardioembolic stroke, large artery atherosclerotic stroke, or cryptogenic/other determined etiology stroke (**Table 3**, **Model 1**). No consistent evidence showed that cumulative mean post-stroke SBP, glucose, or LDL cholesterol attenuated the association between ischemic stroke subtype and post-stroke global cognition (**Table 3**, **Model 2**).

**Table 3.**
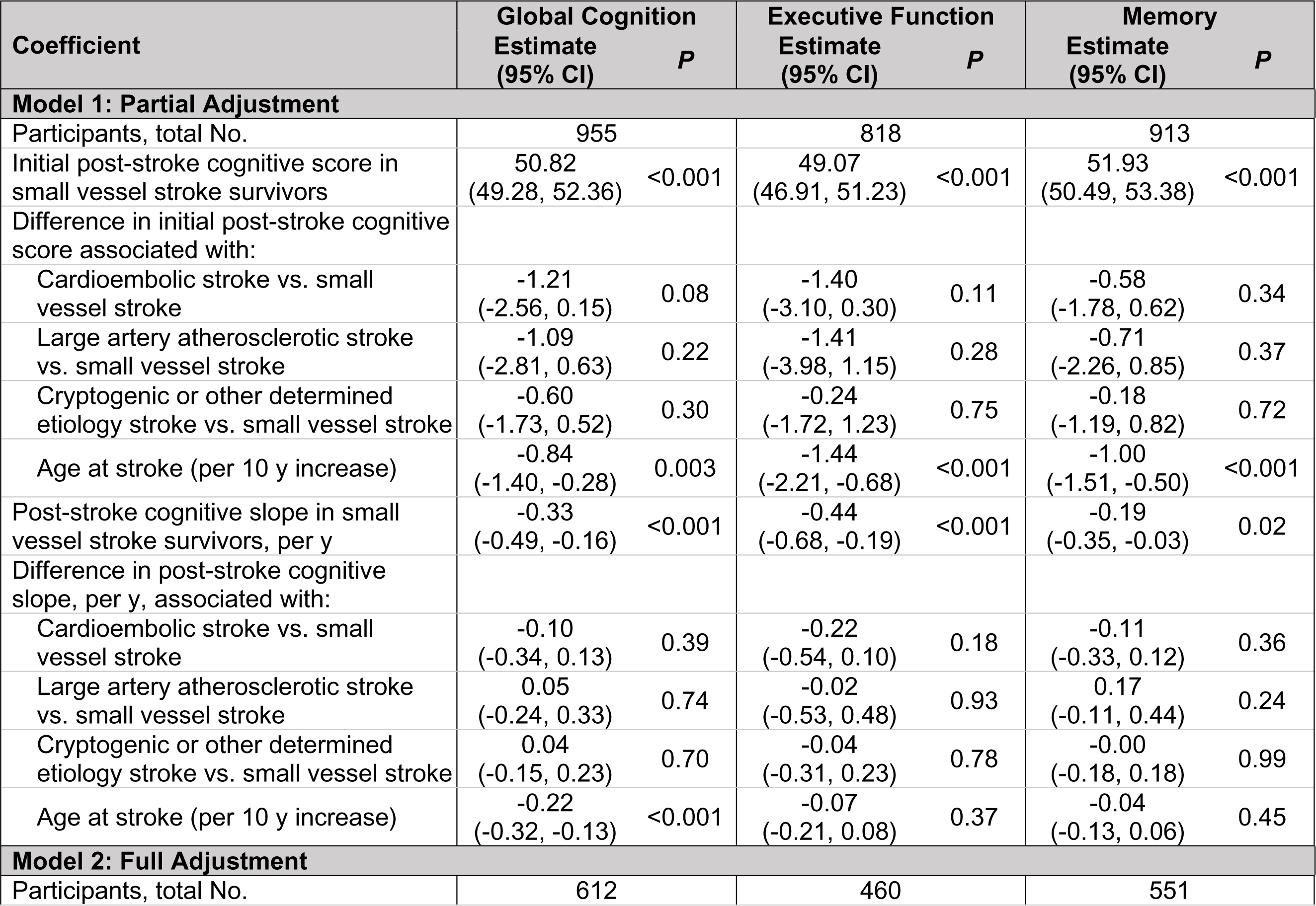
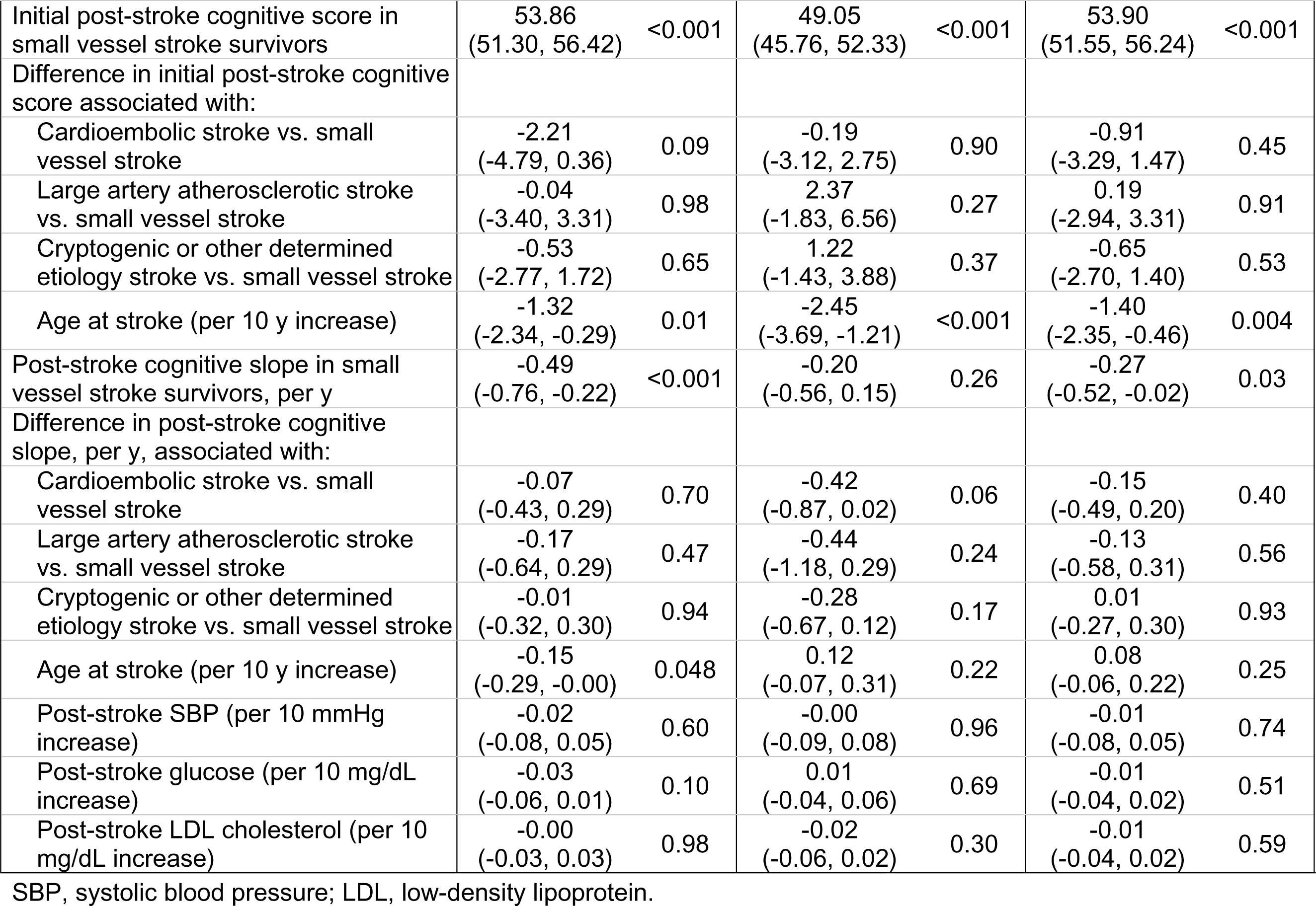
Association of ischemic stroke subtype and post-stroke cognitive decline.

### Changes in Executive Function and Memory

On average, initial post-stroke executive function score was 48.49 (95% CI, 46.83, 50.16) in ischemic stroke survivors and did not differ in hemorrhagic stroke survivors (difference, -0.30 [95% CI, -2.26, 1.66] points; *P*=0.77) after adjusting for demographics and mean pre-stroke cognition (**Table 2**, **Model 1**). On average, initial post-stroke memory score was 52.28 (95% CI, 51.07, 53.49) in ischemic stroke survivors and was lower in hemorrhagic stroke survivors (-1.43 [95% CI, -2.84, -0.02] points; *P*=0.047) after adjusting for demographics and mean pre-stroke cognition (**Table 2**, **Model 1**). This difference was no longer statistically significant after adjustment for cumulative mean post-stroke SBP, glucose, and LDL cholesterol levels (-1.42 [95% CI, -3.80, 0.95] points; *P*=0.24) (**Table 2**, **Model 2**).

On average, ischemic stroke survivors experienced declines in executive function (-0.48 [95% CI, -0.59, -0.36] points/year; *P*<0.001) and memory (-0.27 [95% CI, -0.36, -0.19] points/year; *P*<0.001) over time after adjusting for demographics and mean pre-stroke cognition (**Table 2**, **Model 1**). Post-stroke executive function and memory declines did not differ between hemorrhagic and ischemic stroke survivors (**Table 2**, **Model 1**). Mean post-stroke SBP, glucose, or LDL cholesterol did not consistently attenuate the associations between stroke type and post-stroke declines in executive function and memory (**Table 2**, **Model 2**).

In the subset with non-missing ischemic stroke subtype, on average, small vessel survivors had an initial post-stroke executive function score of 49.07 (95% CI, 46.91, 51.23) and an initial post-stroke memory score of 51.93 (95% CI, 50.49, 53.38; *P*<0.001). Initial executive function and memory score did not differ between small vessel survivors and survivors of cardioembolic stroke, large artery atherosclerotic stroke, or cryptogenic/other determined etiology stroke (**Table 3**, **Model 1**). Small vessel survivors experienced declines in executive function (-0.44 [95% CI, -0.68, -0.19] points/year; *P*<0.001) and memory (-0.19 [95% CI, -0.35, -0.03] points/year; *P*=0.02) over time after adjusting for demographics and mean pre-stroke cognition (**Table 3**, **Model 1**). Post-stroke declines in executive function and memory did not differ between small vessel survivors and survivors of cardioembolic stroke, large artery atherosclerotic stroke, or cryptogenic/other determined etiology stroke (**Table 3**, **Model 1**). No consistent evidence showed that cumulative mean post-stroke SBP, glucose, or LDL cholesterol attenuated the association between ischemic stroke subtype and post-stroke executive function or memory declines (**Table 3**, **Model 2**).

### Sensitivity Analyses

Results were consistent in stroke type analyses adjusting for post-stroke depressive symptoms (**eTable 1**). In the ischemic stroke subtype sample, initial post-stroke global cognition score was significantly lower in cardioembolic stroke survivors compared to small vessel survivors with adjustment for post-stroke depressive symptoms (-1.40 [95% CI, -2.76, -0.04] points/year; *P*=0.04) (**eTable 2**, **Model 1**), although this difference was no longer statistically significant after adjustment for cumulative mean post-stroke VRF levels (-2.16 [95% CI, -4.75, 0.44]; *P*=0.10) (**eTable 2**, **Model 2**). Results were consistent in stroke type and ischemic stroke subtype analyses adjusting for ApoE genotype (**eTables 3** and **4**), excluding participants with history of stroke at baseline (**eTables 5** and **6**), and censoring observations at time of second incident stroke (**eTables 7** and **8**).

## DISCUSSION

In this pooled cohort study with 1,143 stroke survivors, on average, stroke survivors had significant declines in global cognition, executive function, and memory. Cognitive declines did not differ by stroke type or ischemic stroke subtype. Observed post-stroke cognitive declines were not attenuated by adjustment for cumulative mean post-stroke SBP, glucose, and LDL cholesterol.

Our results extend prior research by showing that all stroke types and ischemic stroke subtypes are associated with similar faster declines in global cognition, executive function, and memory, controlling for cumulative mean *post-stroke* VRF levels and pre-stroke cognitive function. In REGARDS (637 ischemic, 55 hemorrhagic), we found that ischemic and hemorrhagic strokes were associated with similar declines in global cognition—measured by the Six-Item Screener—with faster declines for cardioembolic stroke and cryptogenic stroke than other ischemic stroke subtypes after adjusting for *pre-stroke* VRF levels measured once at cohort baseline and pre-stroke cognition; however, we did not include post-stroke VRF levels.^5^ The pooled cohort study’s larger sample size and harmonized cognitive outcomes, including in-person cognitive assessment in multiple cohorts, might have enabled us to detect significant cognitive declines in executive function and memory associated with hemorrhagic stroke and other ischemic stroke subtypes. Our findings are consistent with a study showing that new post-stroke dementia risk is similar between ischemic and hemorrhagic stroke after adjusting for stroke severity and demographics.^2^ Our results also agree with a meta-analysis showing that similar proportions of individuals with lacunar and non-lacunar stroke (16 studies, n=6,478) had mild cognitive impairment or dementia up to four years post-stroke.^24^

No consistent evidence supported our hypothesis that post-stroke VRF levels partially explain relationships between stroke types, ischemic stroke subtypes, and post-stroke cognitive trajectories, with one exception. Hemorrhagic stroke survivors had borderline lower initial post-stroke memory scores than ischemic stroke survivors, consistent with prior research,^5^ but not after adjustment for post-stroke VRF levels. Post-stroke VRF levels might partially explain the relationship between stroke type and initial post-stroke memory scores. Alternatively, they might not, and the change in statistical significance with VRF adjustment might result from a smaller sample size (702 vs. 1,100) as the point estimate for the association stayed the same and the post-stroke VRF levels were not significantly associated with the outcome. More frequent measurements of VRF levels might be needed to detect their effect on post-stroke cognitive trajectories.

These findings suggest that strokes might increase cognitive decline and dementia risks through mechanisms independent of stroke type, ischemic stroke type, and pre-stroke cognitive decline. Potential mechanisms are vascular, neurodegenerative, immune-mediated, inflammatory, and secondary neurodegeneration.^25-29^ In a prior study from the same pooled cohort, higher cumulative mean post-stroke glucose levels were associated with faster decline in global cognition (-0.04 points/year faster per 10 mg/dL increase [95% CI, -0.08, -0.001 points/year]; *P*=0.046) controlling for pre-stroke and post-stroke VRF levels and other covariates. In our study, higher post-stroke glucose levels were not significantly associated with faster post-stroke global cognitive decline, though the effect size was similar (-0.03 points/year faster per 10 mg/dL increase [95% CI, -0.06, 0.01 points/year]; *P*=0.11). Different samples and modeling might explain the differences.

Study strengths are large sample size, inclusion of Black stroke survivors, and population-based sampling, which increases generalizability and is unique for a longitudinal design. Objective measurements of cognition before and after incident stroke enabled us to estimate the association between stroke type and ischemic stroke subtype with cognitive decline, controlling for pre-stroke cognition. Each cohort systematically measured cognitive domains affected by stroke: global cognition, memory, and executive function.

This study has limitations. Information on stroke severity, stroke location, brain imaging, functional status, and other cognitive domains (e.g., language, visuoperception) was unavailable for the analysis. Some cohort study visits included brief rather than extensive neuropsychological assessments, which could reduce sensitivity to subtle cognitive changes. Although linear-effects models do not account for competing risks of death, prior analyses requiring ≥2 post-stroke cognitive assessments yielded similar results.^11^ Selective attrition of cognitively impaired stroke survivors and our assumption that participants’ post-mortem cognitive data are missing at random could underestimate cognitive decline rate. However, the assumption is valid to answer the research question testing differences in post-stroke cognitive trajectories associated with stroke type and ischemic stroke subtype. We used a fixed effect for cohorts because the small number of cohorts precludes estimating random effects for each cohort, potentially producing conservative estimates of cognitive slope differences.

Although our study had a greater percentage of strokes classified as cryptogenic/other determined etiology and a smaller percentage of strokes classified as large-artery atherosclerotic than previous studies, it had a similar percentage of cardioembolic and lacunar/small vessel strokes.^16^ Cryptogenic strokes might include ischemic strokes with limited or incomplete diagnostic evaluation, leading to measurement error and potentially biasing results of differences between ischemic stroke subtypes toward the null. We did not examine incident dementia because REGARDS lacked this information at time of analysis. The number of hemorrhagic strokes was small.

Our results suggest that stroke survivors, especially older stroke survivors, warrant careful long-term monitoring for cognitive decline regardless of stroke type or ischemic stroke subtype. Older age at stroke was associated with faster cognitive decline, supporting that older individuals have higher risk of adverse post-stroke cognitive outcomes. This observation is highly relevant as the increasing life expectancy in the population will result in a larger population at risk of post-stroke cognitive decline and dementia. Research identifying modifiable causes of post-stroke cognitive decline and potential interventions to slow or halt it is needed.

## CONCLUSION

In this pooled cohort study, on average, stroke survivors demonstrated decline in global cognition, executive function, and memory. We found no evidence that post-stroke cognitive declines differed by stroke type or ischemic stroke subtype or that post-stroke VRF levels attenuated associations between stroke type and ischemic stroke subtype with cognitive declines.

## Data Availability

Deidentified participant data if with the approval of a proposal by the individual cohorts and principal investigator and with signed data use agreements between all relevant institutions.

## NON-STANDARD ABBREVIATIONS AND ACRONYMS

VRF: vascular risk factors
SBP: systolic blood pressure
LDL: low-density lipoprotein
ApoE: apolipoprotein E
CES-D: Centers for Epidemiologic Studies Depression Score
ARIC: Atherosclerosis Risk in Communities Study
CHS: Cardiovascular Health Study
FOS: Framingham Offspring Study
REGARDS: Reasons for Geographic and Racial Differences in Stroke Study

## ACKNOWLEDGEMENTS

The content is the authors’ responsibility and does not represent the National Institutes of Health’s (NIH) official views.

## SUPPLEMENTAL MATERIAL

Supplemental eMethods

Supplemental eTables 1-8

References 19, 20, 22, 23, 30

